# Effect of the COVID-19 pandemic on drug-resistant tuberculosis treatment outcomes at a national referral hospital in Sierra Leone, 2017 to 2022: a retrospective study

**DOI:** 10.1101/2025.03.17.25324153

**Authors:** Josephine Amie Koroma, Mariama Mahmoud, Bailah Molleh, Stephen Sevalie, Adrienne K. Chan, Sharmistha Mishra, Sulaiman Lakoh, Joseph Sam Kanu

## Abstract

**Introduction:** Sierra Leone is one of the 30 high TB burden countries, with an incidence rate in 2023 of 286 per 100,000 population. Despite progress in case notification and treatment coverage, around 5,000 cases of TB in Sierra Leone are missing each year. Challenges with notification of drug-susceptible TB can result in drug-resistant TB. The COVID-19 pandemic has further compounded these challenges, resulting in increased drug-resistant TB cases and poor treatment outcomes. This study highlights the effect of COVID-19 on drug-resistant TB treatment outcomes.

**Methods:** We conducted a cross-sectional retrospective analysis of newly identified drug-resistant TB cases in Sierra Leone using data from the national drug-resistant TB database from January 2017 to December 2022, pre-COVID-19, during COVID-19, and post-COVID-19. Data was analysed using STATA. Descriptive analysis was used to summarise the demographic and clinical characteristics and treatment outcomes of drug-resitant TB cases. We used logistic regression to examine the association between time-period and treatment outcomes, after adjusting for age, gender, nutritional status, HIV status and treatment regimens. A p-value of <0.05 was considered statistically significant at 95% confidence level.

**Results:** Of the 701 drug-resistant TB patients, 383 (54.6%) were registered in the pre-COVID-19 period, 228 (32.5%) during COVID-19, and 92 (12.8%) in the post-COVID-19 period. Pre-treatment TB cases reduced from 359 (92.5%) in the pre-COVID-19 period to 80 (30.9%) in the COVID-19 period. New treatment cases increased from 29 (7.5%) to 159 (61.4%) during COVID-19. The proportion of drug-resistant TB that completed treatment decreased from 74.7% in the pre-COVID-19 period to 63.3% during COVID-19 and 68.5% post-COVID-19. There were more cases of successful treatment outcomes in the pre-COVID-19 period (74.7%) than in the COVID-19 period (63.3%). Compared with the pre-COVID-19 period, the odds of a successful outcome were 42% less than in the COVID-19 period (OR 0.58, 95% CI 0.42 to 0.82).

**Conclusion:** We reported a decline in the proportion of drug-resistant TB cases with successful treatment outcomes during COVID-19 and a rapid recovery in the post-COVID-19 period, emphasising the need for robust mitigation strategies for drug-resistant TB management during public health emergencies.

## Background

Tuberculosis (TB) infects 1.2 billion people worldwide each year and caused an estimated 1.25 million deaths in 2023 [1, 2]. Sierra Leone is one of the 30 high TB burden countries, which together account for 87% of the global TB burden. The TB incidence rate in Sierra Leone in 2023 was 273 per 100,000 population [2].

With tremendous progress in case notification by the country’s national TB program in the past ten years, the number of confirmed TB cases increased from 13,195 in 2010 to 17,865 in 2019 [1]. In recent years, these figures have even improved further. Of the 24,000 TB cases expected in 2024, 22,381 (93%) were notified to the National TB Programme. Several challenges in TB care affect sustainability of these gains. The need for prolonged TB treatment increases the risk of treatment interruptions, leading to missed cases and the development of resistance to the most effective TB drugs, such as rifampicin and isoniazid [3, 4].

In 2020, an estimated 101,000 cases of drug-resistant TB were reported in Africa [1]. In Sierra Leone, the prevalence of drug-resitant TB was estimated to be 4.3 per 100,000 population in 2023, including 2.8% of newly diagnosed cases and 21% of retreated patients [2]. This huge burden of drug resistant TB poses additional challenges to TB service delivery in the country.

The early stages of the COVID-19 pandemic portray its adverse impact on adherence to drug-resistnt TB treatment, which was exacerbated by movement restrictions that disrupted health systems and implementation of TB services [5, 6]. Subsequent waves of the pandemic potentially increase TB transmission, delay detection, thereby exacerbating challenges and leading to poorer treatment outcomes and increased drug resistance [7-9].

To date, no studies have evaluated the impact of COVID-19 on treatment outcomes for drug-resistant TB in Sierra Leone. Global evidence is limited to only 6 articles on PubMed using the search term ‘impact of COVID-19 AND treatment outcomes of drug resistance TB’ on October 29, 2024, and none reported data specifically on drug resistance TB treatment outcomes [10-15]. Understanding the impact of COVID-19 on drug-resistant TB treatment outcomes has important policy implications for safeguarding essential health services in future public health emergencies.

This study aims to document the impact of COVID-19 on drug-resistant TB services in Sierra Leone by describing and evaluating the demographics, clinical characteristics, and outcomes of newly registered drug-resistant TB cases before, during, and after COVID-19.

## Methods

### Study Design

We conducted a retrospective cross-sectional study to examine newly registered drug-resistant TB cases between January 1, 2017, and December 31, 2022. Individual patient data were extracted from the National drug-resistant TB database.

### Study Setting

We used data from adults patients (aged 18 years or older) with drug-resistant TB attending the National TB Referral Hospital (Lakka Government Hospital), a 150-bed hospital in western Sierra Leone. The hospital regularly admits patients with drug-sensitive TB and drug-resistant TB for intensive phase treatment as recommended by the national and World Health Organization (WHO) treatment guidelines. After discharge, the patients receive treatment during the continuation phase as outpatient and are closely followed up with medication refills, laboratory tests, and clinical evaluations until they are cured. The hospital has six doctors, 90 nurses, 19 laboratory scientists/technicians, and four radiographers.

In October 2020, Sierra Leone started using a standardized short multi-drug resistant-TB regimen to replace the long conventional regimens. In 2022, bedaquiline, pretomanid, linezolid and moxifloxacin (BPaL-M) and BPaL (if fluoroquinolone-resistant) were adopted to treat multi-drug-resistant TB /rifampicin resistant-TB for all cases aged 14 years or older [16].

### Study Population

Data on drug-resistant TB /rifampicin-resistant (RR)-TB patients were obtained from the National drug-resistant TB Database. A total of 739 cases were registered, but we excluded patients still receiving treatment and patients with missing variables, leaving a total of 701 cases for outcome analysis.

### Data Sources and Variables

Individual deidentified data from the National TB Drug-resistant TB Database were extracted on September 24, 2023. We collected data on age, gender, body mass index (BMI), HIV status, TB treatment status (new or retreatment), treatment regimen (short vs. long) and treatment outcomes {successful (cured, treatment completed) and unsuccessful (death, loss to follow-up, treatment failure and not evaluated)} [17]. TB treatment outcomes were defined in accordance with the WHO reporting framework for patients treated for RR-TB, multi-drug resistant TB and extensive drug-resistant TB using second-line TB medicines (Table 1).

**Table 1:** Outcomes for Rifampicin Resistant-TB/Multi-Drug Resistant-TB patients treated using second-line treatment.

| Outcome | Definition |
| --- | --- |
| Cured | Treatment completed as recommended by the national policy without evidence of failure AND three or more consecutive cultures taken at least 30 days apart are negative after the intensive phase. |
| Treatment complete | Treatment completed as recommended by the national policy without evidence of failure BUT no record that three or more consecutive cultures taken at least 30 days apart are negative after the intensive phase. |
| Treatment failed | Treatment terminated or need for permanent regimen change of at least two anti-TB drugs because of:<br>• lack of conversion by the end of the intensive phase or<br>• bacteriological reversion in the continuation phase after conversion to negative or<br>• evidence of additional acquired resistance to fluoroquinolones or second-line injectable drugs or<br>• adverse drug reactions |
| Died | A patient who dies for any reason during the course of treatment |
| Lost to follow up | A patient whose treatment was interrupted for two consecutive months or more |
| Not evaluated | A patient for whom no treatment outcome is assigned. This includes cases “transferred out” to another treatment unit and whose treatment outcome is unknown |
| Treatment success | The sum of cured and treatment completed |
| Unsuccessful treatment outcome | The sum of death, failure, loss to follow-up, and not evaluated |

### Analysis

Data was analysed using STATA. Descriptive analysis was used to summarise the data the demographic features, clinical characteristics and the treatment outcomes of drug-resitant TB cases using proportion. We used logistic regression to examine the association between time-period and treatment outcomes (successful vs. not successful), after adjusting for potential confounders (age, gender, nutritional status, HIV status and treatment regimens), and we reported crude odds ratios (OR) and adjusted odds ratios (aOR) accordingly. A p-value of <0.05 was considered statistically significant at 95% confidence level.

### Ethics

Ethical approval was obtained from the Sierra Leone Ethics and Scientific Review Committee with approval number 017/05/2023. Routine data was abstracted under a waiver of informed consent from the ethics committee and approval from the health facility management. The authors did not have access to information that could identify individual participants during or after data collection.

## Results

### Demographic and clinical profile of registered patients with drug-resistant TB

Of the 701 drug-resistant TB patients, 383 (54.6%) were registered in the pre-COVID-19 period, 228 (32.5%) during COVID-19, and 92 (12.8%) in the post-COVID-19 period. Nearly three-fourths (284, 73.2%) of patients were aged 15 to 44 years. Males comprised a larger proportion (523, 68.5%) of patients with drug-resistant TB in all periods. However, the proportion of women with drug resistant TB increased from 25.8% in the pre-COVID-19 period to 33.6% during COVID-19 and 31.5% in the post-COVID-19 period. The re-treatment TB cases reduced from 359 (92.5%) in the pre-COVID-19 period to 80 (30.9%) in the COVID-19 era. On the other hand, new treatment cases increased from 29 (7.5%) to 159 (61.4%) during the COVID-19 period (Table 2). Malnutrition among drug resistant TB patients increased from 65.7% in the pre-COVID-19 period to 70.7% in the COVID-19 era. HIV positivity among drug-resistant TB cases increased from 17.5% in the pre-COVID-19 period to 26.6% during COVID-19 and 21.7% in the COVID-19 period (Table 2).

**Table 2.**
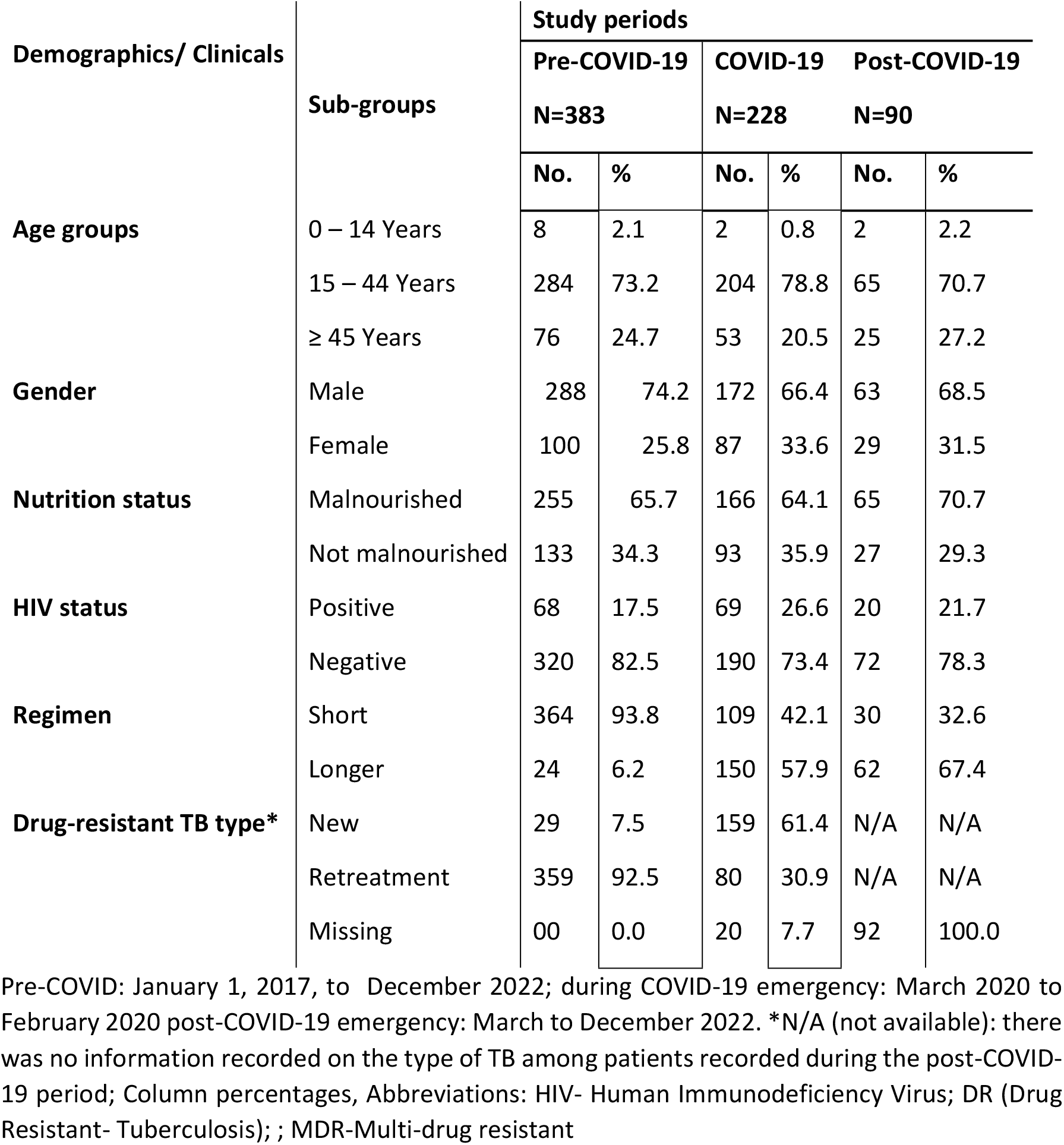
Demographic and clinical profile of registered patients with drug-resistant TB.

| Demographics/ Clinicals | Sub-groups | Study periods |  |  |  |  |  |
| --- | --- | --- | --- | --- | --- | --- | --- |
|  |  | Pre-COVID-19 |  | COVID-19 |  | Post-COVID-19 |  |
|  |  | N=383 |  | N=228 |  | N=90 |  |
|  |  | No. | % | No. | % | No. | % |
| Age groups | 0 – 14 Years | 8 | 2.1 | 2 | 0.8 | 2 | 2.2 |
|  | 15 – 44 Years | 284 | 73.2 | 204 | 78.8 | 65 | 70.7 |
|  | ≥ 45 Years | 76 | 24.7 | 53 | 20.5 | 25 | 27.2 |
| Gender | Male | 288 | 74.2 | 172 | 66.4 | 63 | 68.5 |
|  | Female | 100 | 25.8 | 87 | 33.6 | 29 | 31.5 |
| Nutrition status | Malnourished | 255 | 65.7 | 166 | 64.1 | 65 | 70.7 |
|  | Not malnourished | 133 | 34.3 | 93 | 35.9 | 27 | 29.3 |
| HIV status | Positive | 68 | 17.5 | 69 | 26.6 | 20 | 21.7 |
|  | Negative | 320 | 82.5 | 190 | 73.4 | 72 | 78.3 |
| Regimen | Short | 364 | 93.8 | 109 | 42.1 | 30 | 32.6 |
|  | Longer | 24 | 6.2 | 150 | 57.9 | 62 | 67.4 |
| Drug-resistant TB type* | New | 29 | 7.5 | 159 | 61.4 | N/A | N/A |
|  | Retreatment | 359 | 92.5 | 80 | 30.9 | N/A | N/A |
|  | Missing | 00 | 0.0 | 20 | 7.7 | 92 | 100.0 |

Pre-COVID: January 1, 2017, to December 2022; during COVID-19 emergency: March 2020 to February 2020 post-COVID-19 emergency: March to December 2022. \*N/A (not available): there was no information recorded on the type of TB among patients recorded during the post-COVID- 19 period; Column percentages, Abbreviations: HIV- Human Immunodeficiency Virus; DR (Drug Resistant- Tuberculosis); ; MDR-Multi-drug resistant

### Treatment outcomes of patients with drug-resistant TB

Treatment outcomes differed across the study periods. The proportion of patients with drug-resistant TB who completed treatment decreased from 74.7% in the pre-COVID-19 period to 63.3% during COVID-19 and 68.5% post-COVID-19. The number of deaths among patients with drug-resistant TB was higher during the COVID-19 period (22.8%) and post-COVID period (20.0%) than in the pre-COVID period (16.7%). Similarly, more patients with drug-resistant TB were lost follow up in the post-COVID-19 peroid (5.6%) and COVID-19 peroid (4.4%) than in the pre-COVID-19 peroid (4.2%). There were more patients with successful treatment outcomes before the COVID-19 pandemic (74.7%) than during COVID-19 (63.3%) and after COVID-19 period (70.0%) (Table 3).

**Table 3.** Clinical characteristics and the treatment outcomes of newly registered drug-resitant TB cases pre-COVID-19, COVID-19 and post-COVID-19.

| Treatment outcomes | Sub-groups | Study periods |  |  |  |  |  |
| --- | --- | --- | --- | --- | --- | --- | --- |
|  |  | Pre-Covid<br>N=383 |  | Covid<br>N=228 |  | Post-Covid<br>N=90 |  |
|  |  | No. | % | No. | % | No. | % |
| **All treatment outcomes** | Complete | 290 | 75.7 | 164 | 71.9 | 63 | 70.0 |
|  | Died | 64 | 16.7 | 52 | 22.8 | 18 | 20.0 |
|  | Failed | 13 | 3.4 | 2 | 0.9 | 4 | 4.4 |
|  | LTFU | 16 | 4.2 | 10 | 4.4 | 5 | 5.6 |
| **Outcomes (Binary)** | Successful | 290 | 75.7 | 164 | 71.9 | 63 | 70.0 |
|  | Unsuccessful | 93 | 24.3 | 64 | 28.1 | 27 | 30.0 |

\*Unsuccessful treatment outcome includes death, failure, and loss to follow-up (LTFU). It does not include “on treatment”.

### Factors associated with successful drug-resistance TB treatment outcomes

Compared with the pre-COVID-19 period, the odds of a successful outcome were 42% less than in the COVID-19 period (OR 0.58, 95% CI 0.42 to 0.82) during the COVID period. The odds of a successful outcome were similar between the pre- and post-COVID periods (OR 0.73, 95%CI 0.45 to 1.21). However, after adjusting for potential confounders (age, gender, nutritional status, HIV status and treatment regimens), there was no significant effect of COVID-19 on drug-resistant TB treatment outcomes (aOR: COVID-19 =0.87(95% CI: 0.57 – 1.33); post-COVID-19=1.26 (95% CI: 0.71 – 2.26).

## Discussion

The study provides a comprehensive assessment of the impact of the COVID-19 pandemic on drug-resistant TB services in Sierra Leone, focusing on demographic and clinical characteristics and treatment outcomes of newly registered drug-resistant TB cases across the different pandemic periods. Utilizing programmatic data, the study reported a significant effect of COVID-19 on drug-resistant TB notifications, and treatment outcomes. There were more cases of drug-resistant TB during the COVID-19 pandemic than before the pandemic. Second, before the COVID-19 pandemic, 74.7% of drug resistant TB cases were successfully treated, compared to 63.3% during COVID-19. There were gender and age disparities, with more cases of drug-resistant TB among young male population. Malnutrition and HIV positivity among drug resistant TB patients increased following the COVID-19 pandemic. Finally, the odds of a successful treatment outcome during COVID-19 were 42% less than in the COVID period.

The marked increase in the number of notified drug-resistant TB cases, particularly among people aged 15 to 44 years, may be explained by the living conditions of young people in poor, overcrowded housing facilities with limited access to healthcare and increase their risk of contracting TB [18]. Movement restrictions and lockdowns during the COVID-19 pandemic may have exacerbated overcrowding and increased the risk of TB infection. In Iraq, there was a significant increase in drug-resistant TB cases among young people aged 15-34 years, similar to our findings [19]. Young people engage in many activities, such as smoking, and have inherent immunopathology and hormonal changes that put them at risk for TB infection [19].

There has been a shift in the gender distribution trend, with an increase in the proportion of male drug-resistant TB cases during the pandemic period. These demographic shifts may underscore the pandemic’s impact on disease surveillance and patient demographics [20] and call for mainstreaming of gender-related interventions in public health emergency preparedness and response in low-income countries.

COVID-19 disrupted health services delivery and supply chain of essential health commodities, leading to poor health outcomes [21]. This study highlighted adverse changes in clinical parameters during the pandemic. The recovery phase of the COVID-19 pandemic saw a notable rise in malnutrition among drug-resistant TB patients compared to pre-pandemic levels. Malnutrition is a public health problem in Sierra Leone [22]. The interaction of social factors such as poverty with COVID-19 and TB may explain the worsening malnutrition among patients with drug-resistant TB in this study. Poverty worsens during the COVID-19 and post-COVID-19 period [23]. Furthermore, restrictions during the pandemic reduced the quality and productivity of plants, meat and eggs, which worsens malnutrition [24].

There was an increased proportion of HIV-positive cases with drug-resistant TB in the post-pandemic period compared to the pre-pandemic period. This finding reflects the higher likelihood of a positive HIV test during the COVID-19 pandemic observed in a previous study in Sierra Leone [25]. The reason for the increased burden of HIV during and after the pandemic could be due to late presentation for care during and after the pandemic [26].

The COVID-19 period also witnessed an uptick in new TB cases and fluctuations in treatment regimens, reflecting the healthcare system’s adaptive responses and challenges during crisis periods [1, 2].

Other consequences of the COVID-19 restriction measures are the adverse effects on TB treatment outcomes. Treatment outcomes exhibited variability across the study periods, with a decline in treatment completion rates, increased treatment failures, and a high proportion of deaths observed during the phase of COVID-19. A similar study done on treatment outcomes in the country indicated that the lockdown imposed by the authorities on mobility and strikes by healthcare workers contributed to missed drug doses, leading to treatment failures and possible deaths [9]. Although there was a decrease in successful treatment outcomes during COVID-19 compared to pre-pandemic levels, a gradual recovery was noted in the COVID-19 recovery period, reflecting on the resilient nature of the health system.

The odds of a successful treatment outcome were significantly lower during the COVID-19 period compared to the pre-COVID-19 period. However, after adjusting for potential confounders such as age, gender, nutritional status, HIV status, and treatment regimens, the study found no significant effect of COVID-19 on drug-resistant TB treatment outcomes. This may suggest that while there were observable impacts on initial treatment success rates, these effects were mitigated by considering broader clinical factors.

Our study has limitations. The study’s analysis of retreatment cases in 2022 was limited by missing clinical variables, potentially influencing the findings. This could compromise the interpretation of the results, necessitating future research to ensure the completeness and accuracy of clinical data. Future research should prioritize data integrity for a better understanding of pandemic dynamics and optimizing outcomes for drug-resistant TB patients in low-income countries.

## Conclusion

We reported a decline in the proportion of drug-resistant TB cases with successful treatment outcomes during the COVID-19 period and a rapid recovery in the post-COVID-19 period, emphasizing the need for robust mitigation strategies for drug-resistant TB management during public health emergencies. Furthermore, our study reported worsening malnutrition and HIV burden among drug-resistant TB patients during the COVID-19 period. This finding highlights the importance of comprehensive patient management strategies for drug-resistant TB to improve healthcare delivery and patient outcomes.

## Data Availability

The datasets used and/or analyzed (10.6084/m9.figshare.28583720) for this study are available from: https://figshare.com/account/items/28583720/edit.

## Acknowledgements

This research was conducted through a partnership between Sustainable Health Systems Sierra Leone, and the Li Ka Shing Research Institute, Unity Health, University of Toronto, Canada. The training model used that resulted in this publication was adapted from the Structured Operational Research and Training Initiative (SORT IT), a global partnership led by the Special Programme for Research and Training in Tropical Diseases at the World Health Organization (WHO/TDR, Geneva, Switzerland), for which SL, AKC and SM are SORT-IT Mentors. Mentorship was provided by Sustainable Health Systems, (Freetown, Sierra Leone); Ministry of Health, Government of Sierra Leone; University of Toronto (Toronto, Canada); Partners in Health Sierra Leone (Koidu, Sierra Leone). The authors would like to acknowledge Kristy Cheuk Yin Yiu and Bailah Molleh for Project Management and Coordination.

## Funding

This research was funded by the Canadian Institutes for Health Research (CIHR) through an Operating Grant (WI1-179883) Addressing the Health Impacts of COVID-19. SM is funded by a Tier 2 Canada Research Chair in Mathematical Modeling and Program Science (CRC File Number 950-232643). The funders had no role in the study design, data collection and analysis, decision to publish, or preparation of the manuscript.

## Conflict of interest

None to declare

## Author’s contribution

Conceptualization and study design: JAK, MM, JSK, AKC, SM, SS, SL

Data curation and validation: JSK, JAK, BM

Methodology and formal analysis: JAK, AKC, SM, JSK

Funding acquisition, resources, supervision: SS, SL, SM, AKC

Writing – original draft preparation: JAK, AKC, JSK, SL, SM

Writing – review & editing: SL, AKC and SM

## Notes

### Competing Interest Statement

The authors have declared no competing interest.

